# Transforming Urban Primary Health Care Through Digitalization: The Sanjeevani Clinic Experience in Madhya Pradesh

**DOI:** 10.64898/2026.01.13.26343577

**Authors:** Tavpritesh Sethi, Jasmine Kaur, Chayan Roychoudhury, Pradeep Singh, Priyanka Das, Manish Singh, Shalini Kapoor, Savita Sharma, Ambey Srivastava, Akhilesh Yadav, Rakesh Kumar

## Abstract

**Problem:** Rapid and unplanned urbanization in LMICs exacerbates health inequities, particularly among the urban poor living in slums. In Madhya Pradesh (MP), India, the state government introduced Sanjeevani clinics to provide free, quality healthcare to these populations. This study highlights lessons from digitizing these clinics and making them AI-ready with an open-source AI framework.

**Approach:** A multisectoral collaboration between the state government, implementation, and academic partners led to their successful digitization and AI-readiness of Sanjeevani clinics. An innovative, three-tablet task-shifting model and Smart Clinic Application reduced the data entry burden. *ChikitsaChakra*, an open-source, probabilistic decision support framework with a voice interface was developed to glean insights from data.

**Local setting:** Madhya Pradesh has a population of over 84 million, with approximately 24 million (29%) living in urban settings. Of these, 6.7 million (28%) resided in urban weak economic clusters. Prior to the introduction of Sanjeevani clinics, the population was heavily reliant on private healthcare, incurring significant out-of-pocket expenditures (OOPE).

**Relevant changes:** Digitization created an evidence base for over 2.6 million primary care consultations, saving over $33 million in cumulative OOPE while also revealing patterns in antibiotic prescribing.

**Lessons learned:** The Sanjeevani clinics” success hinged on multisectoral collaboration among government bodies, nonprofits, academia, and private partners, crucial for digitalization and AI-readiness. The innovative three-tablet model and application improved data quality, reducing healthcare workers” burdens and enhancing monitoring and evaluation. *ChikitsaChakra* provided essential insights.

## INTRODUCTION

The world has become predominantly urban, with urbanization continuing to accelerate globally. According to the United Nations, 55% of the world”s population resided in urban areas as of 2018, a proportion that is expected to rise to 68% by 2050(1). In LMICs, where infrastructure development often struggles to keep pace with population growth, this rapid and uncontrolled urbanization has introduced new healthcare challenges and equity issues(2). As India progresses as one of the fastest-growing economies, nearly 40% of its population is projected to be urban by 2036(3). With a current shortfall of 39% for primary healthcare for urban areas in India(4), the urban healthcare infrastructure is strained, pushing the urban poor into overcrowded and under-resourced environments. Further, evidence on gender-based inequities in healthcare access is limited. Studies suggest men often travel farther than women to seek care, yet comprehensive data on primary healthcare access remains lacking(5). Understanding these disparities is essential, as inequitable access to affordable healthcare has significant economic consequences. The National Sample Survey (2017-18) reported that for each consultation, patients spent an average of $4.1 for outpatient services in public health facilities, $8.5 for private doctors, and $13 in private hospitals(6). Thus, the affordability of private healthcare is a larger challenge, leaving the urban poor vulnerable to catastrophic health expenditure (CHE)(7). Nearly 584 million consultations per year and around 266 consultations per day per urban PHC are required for the management of non-communicable diseases (NCDs)(3,4,9).

The Ayushman Bharat Digital Mission (ABDM) in India represents a transformative initiative to enhance healthcare accessibility and equity. Launched by the Government of India, ABDM establishes a critical framework for digital health records and the longitudinal recording of health information with quality data as a cornerstone. Here we report learnings from implementing the Smart Clinic Application, a digital intervention designed to align seamlessly with ABDM”s objectives of strengthening primary healthcare delivery, interoperability, data quality, and privacy. This application contributes to a more efficient healthcare system by streamlining data entry and ensuring high-quality electronic health records, a cornerstone of ABDM. By providing comprehensive, accurate patient data, it enables doctors to make well-informed decisions and enhances coordinated care through seamless information sharing within clinics. The application was designed with a focus on interoperable standards such as SNOMED-CT for ensuring data quality. Additionally, the application”s real-time data analytics support optimized resource planning and enable AI-driven predictive modeling for supply chains, resonating with ABDM”s emphasis on data-driven decision-making. We also discuss the need to implement ABDM”s Health ID to achieve a continuum of care and analytics.

### LOCAL SETTING

Madhya Pradesh (MP), the second-largest state by area in India has an urban population of around 24 million(8), of which approximately 6.7 million urban population resided in urban weak economic clusters. Until 2021, 35%(9) of households from lower wealth quintiles preferred private health facilities for seeking healthcare. Prior to the introduction of Sanjeevani clinics, more than 58% relied primarily on personal savings and incurred an average out-of-pocket medical expenditure of $3 and $35 per treated spell of ailment in non-hospitalized public and private settings respectively(10). AI models were developed and piloted on data from Bhopal district in MP.

## APPROACH

Recognizing these challenges, the Madhya Pradesh state government introduced Sanjeevani Clinics to provide access to primary care at the doorstep(11) and subsequently digitized them. National Health Mission of Madhya Pradesh repurposed the existing community spaces for the Sanjeevani clinics through inter department collaboration between NHM and Urban Administration and Housing Department. Between 2020 and 2023, the Wadhwani Initiative for Sustainable Healthcare (WISH) provided technical support to the MP government for digitization of healthcare services in alignment with the principles of ABDM to achieve scalability, equity, efficiency, and data-driven decision-making (12).

Digitized and comprehensive data collection and analytics for each facility were done through the introduction of a complex intervention achieved through multisectoral collaboration. The Smart Clinic Application (SCA) implemented as an innovative three-tablet data entry model (Figure 1a) was introduced by the WISH Foundation to achieve efficient task-shifting between doctors, nurses, and pharmacists. The smart clinic application comprises three primary modules, each tailored to streamline specific workflows and enhance efficiency across clinical roles. The Nurse”s Tablet module assists nurses in patient registration, recording vital signs and medical history, managing patient queues, and supporting doctors with data entry and patient care tasks. The Doctor”s Tablet module is designed for physicians, allowing them to access comprehensive patient history, record new diagnoses, prescribe medications, order lab tests, and analyze patient data in real-time. Physicians can generate digital prescriptions, which are instantly available on the Pharmacist”s Tablet, facilitating seamless pharmacy operations. The pharmacist module enables the receipt of digital prescriptions, inventory management, medication counseling, and billing, ensuring efficient dispensing and payment processes. Data security and privacy were ensured through role-based access for each module ensuring only relevant information made visible in each module. All patient records were encrypted using SHA 256 encryption, and only anonymized data for Bhopal district was shared for analytics and modeling.

**Figure 1.**
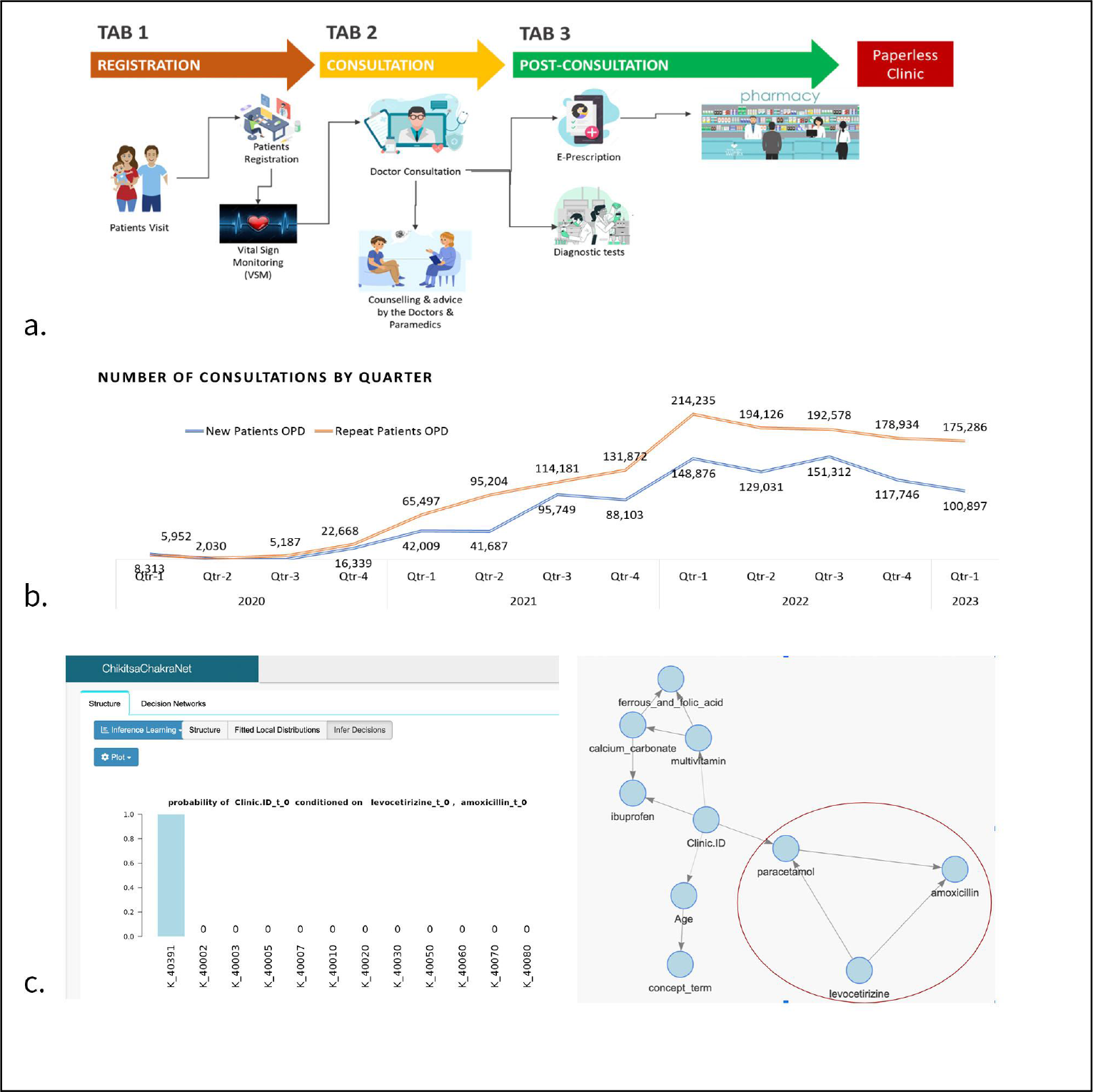
**a. Three-tablet data entry model**. Implemented as the Smart Clinic Application across 23 Districts of Madhya Pradesh, it achieved task-shifting and competency-based training leading to better data quality. **b. Increasing quarterly consultations**. A quarterly increase of 30,000 patients (p<0.001, R^2^ = 0.81) (not shown) and repeat visits quarterly increase of 19,000 patients ((p<0.001, R^2^ = 0.87) was observed. **c. Evidence-based and AI-ready advocacy**. The Dynamic Bayesian Network model developed for the Bhopal district showed clinic and doctor-specific associations between antibiotics, anti-allergic and anti-pyretic medications highlighting the need for data-informed and localized advocacy for rational use of drug combinations. The open web-application and generalizable framework is available at https://github.com/tavlab-iiitd/ChikitsaChakra.

Comprehensive training, including a specific module on the three-tablet system, along with comprehensive primary health care and its delivery mechanisms in line with the health wellness center approach was provided to all clinic staff. Incentive-based salaries were provided to the medical officers administering services in these clinics. 120 essential drugs and 63 types of diagnostic tests based upon Government of India guidelines for Health and Wellness Centres (HWCs) were sourced using a Public-Private-Partnership model between NHM MP and local private providers. In addition to providing clinical services, each clinic was assigned Outreach Nurses (ANMs) responsible for health, nutrition, and Water, Sanitation and Hygiene (WASH) activities for approximately 10,000 residents, ensuring community engagement and proactive health management.

An academic partner, Indraprastha Institute of Information Technology (IIIT-Delhi) provided support for the development of an artificial intelligence (AI) framework to make these Sanjeevani clinics in Bhopal district AI-ready with a novel, open-source, generalizable Dynamic Bayesian Network model to understand spatio-temporal prescription patterns. The core innovation of *ChikitsaChakra* was in the Dynamic Bayesian Network model learned using *R Shiny*. The interactive voice and visual interface, ***ChikitsaChakra***, was developed using R Shiny and GPT-3-based Whisper frameworks, aimed at enhancing accessibility for healthcare providers, particularly those with limited digital experience. Although the Dynamic Bayesian Network model was able to capture the prescription patterns, the voice-based interface for the model did not achieve the desired accuracy of translating voice queries to model predictions. However, voice-based models are important to make it more accessible to end users and future direction involves integrating newer models for this task. The open web application and generalizable framework is available at https://github.com/tavlab-iiitd/ChikitsaChakra.

## RELEVANT CHANGES

Launched in December 2019, the Sanjeevani initiative rapidly scaled to meet the growing demands for primary healthcare services, driven by strategic implementations and innovative use of technology. Funded by the state government, the cost of implementing the three-tab model was approximately USD 1753 per clinic with operation costs of approximately USD 22,577 per annum per clinic. Growing from five to 110 clinics across twenty-four districts by December 2021, over one million digitized consultations have been provided through these clinics by the end of 2021. Data from these clinics showed that 2,613,607 beneficiaries accessed services, showing a significant increase in service utilization from 35,000 in the first quarter of 2020 to 289,753 in the first quarter of 2023, an average quarterly increase of 30,000 patients (p<0.001, R^2^ = 0.81). Repeat visits increased by 19,000 patients (p<0.001, R^2^ = 0.87) every quarter. Notably, services for pregnant women, including the identification of high-risk cases, were significantly expanded, with nearly 9% of them identified as high-risk and appropriately referred. The success of this model has led the government to propose 600 new clinics across all 51 districts with the help of the XV Finance Commission budget.

Before the initiation of Sanjeevani clinics, about 35% of lower wealth quintile urban households preferred private facilities due to the lack of nearby public health options(8), spending approximately $35 per visit(9). Sanjeevani clinics served 2.6 million OPDs in 23 districts of Madhya Pradesh between 2020 to 2023. Over the years, more females accessed Sanjeevani Clinic services in the Bhopal district, suggesting greater equity in access to this model compared to previous reports(5). However, without the implementation of the ABHA ID, it was not possible to analyze patient transitions between Sanjeevani Clinics and telemedicine services. This highlights the importance of ABHA ID in facilitating a better understanding of the continuum of care.

Digitization of data allowed for continuous monitoring, availability of improved quality of data, accountability and evaluation. Although direct evidence was not available, the three-tablet model significantly improved data quality by aligning training with user competency and alleviating the data-entry burden. Indirect evidence indicates that, without this model, there is often a misalignment between the trained personnel and the end-users of the application, resulting in inconsistencies and reduced data integrity.

Making Sanjeevani Clinics AI-ready with the development of the open-source *ChikitsaChakra* framework represents an early step towards data-driven, evidence based and collaborative decision-making in primary care settings. The explainable and interactive Dynamic Bayesian Network revealed important insights such as the need to inform antibiotic usage at the primary care level to provide personalized guidelines at the doctor, clinic and district levels. However, while AI-based voice interaction with the model shows promise, it was deemed unsuitable for roll-out due to the need for further improvements in voice model accuracy.

## LESSONS LEARNED

Our experience with digitizing Sanjeevani Clinics and developing data-driven insights provides valuable guidance on best practices and potential pitfalls in using digitization in pursuit of Universal Health Coverage (UHC) in India. As per our knowledge, this is the first example of digitalisation of urban community clinics in India through a low-cost smart clinic application involving three tabs achieving task shifting. A similar implementation of the three-tab model was also carried out as a part of WISH”s support to Mohalla clinics in Delhi. This effort is aligned with the robust digital health strategy that has evolved in India since the introduction of the National Health Policy of 2017 aimed at expanding access to quality care, lowering healthcare costs, and integrating digital technology to streamline service delivery. However, significant barriers such as limited digital infrastructure, lack of digital literacy, data privacy concerns, and disparities in internet access continued to pose challenges for such initiatives. Before the introduction of the Smart Clinic Application, Sanjeevani Clinics faced significant challenges in delivering quality care and achieving continuity of care due to their reliance on paper-based systems. Accountability for data entry and efficient task shifting were critical challenges for capturing high-quality, analyzable data. The three-tab SCA application combined with interoperable standards and robust data protection framework established the foundation for data-driven, evidence-based insights to support equitable population health in Madhya Pradesh. The three-tablet SCA model and *ChikitsaChakra* framework are generalizable to primary care settings across India and LMICs. However, both implementation models and AI models need to be contextualized for local settings based on availability of human resources and prevalent healthcare patterns. Government leadership, strategic use of community spaces and multisectoral coordination was critical for achieving digitization and AI-readiness of the Sanjeevani clinics. For this complex digital intervention, the separate ongoing evaluation(13) of digitization model and AI frameworks have already provided valuable insights to guide urban primary health care in India and other LMICs. Improvement in data quality was an unintended positive outcome of the distributed data entry approach. For new patients, nurses filled 24 fields, doctors entered 11 fields, and pharmacists entered 2 fields. For repeat patients, these were reduced to 9 fields for nurses, 4 fields for doctors, and 2 fields for pharmacists.

However, we identified a compliance gap in the effective use of SNOMED-CT based drop-down menus. Health care workers frequently opted for the unstructured text field “Other” instead of selecting appropriate menu items, leading to persistent data quality issues that required retrospective cleaning. Another source of potential error was the absence of a universal Health ID. Although aggregate medication use remained unaffected, duplicated patient IDs created confusion, hindering individualized recommendations in primary care. Effective implementation of the national Ayushman Bharat Health Account (ABHA) will be pivotal in addressing this challenge and for achieving continuum-of-care. Our experience with generating insights from the large volumes of digitized data through the use of *ChikitsaChakra* framework highlighted the need for targeted advocacy for rational use of drug combinations. However, it is also highlighted for continuous and iterative improvement of user design to make this framework more accessible for generating insights at the level of policymaker and medical officer. Better interfaces such as voice-based interaction with the model are needed and are available as an experimental feature in the *ChikitsaChakra* framework.

The success of the Sanjeevani clinics and the ChikitsaChakra framework underscores the potential for scaling similar digital health interventions across India and other LMICs. To generalize and extend these models, several key actions are needed:

1. **Implementation of Universal Health IDs**: Introducing universal Health IDs is critical for streamlined patient tracking, ensuring continuity of care, and enabling personalized medical recommendations.
2. **Enhanced Training and Compliance**: Strengthening training programs and compliance with SNOMED-CT standards will improve data quality and facilitate more effective use of health information systems.
3. **Improved User Interfaces**: Developing more intuitive user interfaces, including voice-based interactions, can enhance usability for healthcare providers, ensuring better data entry and retrieval processes.
4. **Continuous Evaluation and Adaptation**: Regular assessments and iterative improvements of the digital health models are essential to adapt to varying local contexts and healthcare needs.
5. **Multisectoral Collaboration**: Sustained collaboration between government bodies, healthcare providers, and technology partners will be crucial for the successful implementation and scaling of these interventions.

The Government of India”s strong emphasis on digital health, coupled with dedicated funding for improving such systems, is solidifying such efforts for long-term sustainability. State Governments, recognizing the potential of Sanjeevani clinics, have allocated significant budget provisions under the PIP to facilitate their expansion. By focusing on these areas, other regions in India and LMICs can adopt and adapt these innovative approaches to enhance primary healthcare delivery, ultimately improving health outcomes and reducing disparities.

### Box 1.

**Summary of main lessons learnt**

1. **Multisectoral Collaboration:** The successful implementation of the Sanjeevani clinics relied on strong collaboration between government bodies, not for profit organizations, academic institutions, and private partners. This collaboration was crucial for achieving the digitization and AI-readiness of healthcare services.
2. **Data Quality Improvement:** The innovative three-tablet model and SCA application for data entry not only significantly improved the quality of data collected but also reduced the data entry burden on healthcare workers freeing up time for quality service provision. This enhancement in data quality facilitated better monitoring and evaluation of healthcare services.
3. **AI-Driven Insights:** The development of the *ChikitsaChakra* framework provided valuable AI-driven insights, such as the need for rational use of antibiotics. These insights are essential for making data-informed decisions and advocating for better healthcare practices at the primary care level.

## Data Availability

The data is not publicly available

## Ethics Statement

The retrospective analysis of these data were conducted following ethical approvals received from the Institutional Review Board of Institute of Health Management Research (IIHMR), Bangalore (IRB Application No. IRB_IHMRB_24Q1_03)

## Acknowledgements

This study was supported by the Alliance for Health Policy and Systems Research (Alliance). The Alliance is able to conduct its work thanks to the commitment and support from a variety of funders. These include Bill & Melinda Gates Foundation, which contributed designated funding and support for this project, along with the Alliance”s long-term core contributors from national governments and international institutions. For the full list of Alliance donors, please visit: https://ahpsr.who.int/about-us/funders. We acknowledge the support of Madhya Pradesh State Government for their leadership and engagement and support of Prashant NS from Institute of Public Health, Bangalore. We acknowledge Ms. Prashasti Agarwal, Ms. Aastha Sharma and Mr. Madhava Krishna for their contributions to the development of Dynamic Bayesian Network framework and Dr Amit Sachan for his support in analyzing Sanjeevani data.

